# Apnea-hypopnea index estimation with wrist-worn photoplethysmography

**DOI:** 10.64898/2026.04.08.26350411

**Authors:** Pedro Fonseca, Marco Ross, Fokke van Meulen, Jerryll Asin, Merel M. van Gilst, Sebastiaan Overeem

## Abstract

**Objective:** Long term monitoring of obstructive sleep apnea (OSA) severity may be relevant for several clinical applications. We developed a method for estimating the apnea-hypopnea index (AHI) using wrist-worn, reflective photoplethysmography (PPG).

**Approach:** A neural network was developed to detect respiratory events using PPG and PPG-derived sleep stages as input. The development database encompassed retrospective data from three polysomnographic datasets (N=3111), including a dataset with concurrent reflective PPG recordings from a wrist-worn device (N=969). The model was pre-trained with (transmissive) finger-PPG signals from all overnight recordings and then fine-tuned to wrist-PPG characteristics using transfer learning. Validation was performed on the test portion of the development set and on a fourth, external hold-out dataset containing both wrist-PPG and PSG data (N=171). Performance was evaluated in terms of AHI estimation accuracy and OSA severity classification.

**Main Results:** The fine-tuned wrist-PPG model demonstrated strong agreement with the PSG-derived gold-standard AHI, achieving intra-class correlation coefficients of 0.87 in the test portion of the development set and 0.91 in the external hold-out validation set. Diagnostic performance was high, with accuracies above 80% for all severity thresholds.

**Significance:** The study highlights the potential of reflective PPG-based AHI estimation, achieving high estimation performance in comparison with PSG. These measurements can be performed with relatively comfortable sensors integrated in convenient wrist-worn wearables, enabling long-term assessment of sleep disordered breathing, both in a diagnostic phase, and during therapy follow-up.

## 1. Introduction

The diagnostic assessment of obstructive sleep apnea (OSA) traditionally relies on polysomnography (PSG), which combines the measurement of breathing with airflow, respiratory effort, and oxygen saturation, as well as sensors to directly assess sleep based on neurological activity, including electroencephalography. The clinical diagnosis of OSA is then established based on the clinical presentation combined with a measure of frequency of respiratory events. The latter is typically quantified by the apnea-hypopnea index (AHI), defined as the number of sleep apneas and hypopneas, divided by the total sleep time [1].

The scoring of respiratory events relies mainly on the analysis of respiratory activity. Consequently, full PSG recordings have also been simplified into smaller polygraphic setups typically restricted to the measurement of airflow (nasal pressure cannulas, oronasal thermistors), peripheral blood oxygen saturation, and respiratory effort (thoracic and/or abdominal respiratory inductance plethysmography (RIP) belts). These polygraphic setups, often referred to as home sleep apnea tests (HSATs), are increasingly used in the diagnosis of OSA. However, they are still obtrusive and uncomfortable due to multiple attached sensors, restricting usage to one or two nights. In addition, due to the lack of objective sleep measurements, these systems use total recording time, rather than total sleep time to estimate the AHI, which can lead to a systematic underestimation of OSA severity [2,3].

These HSAT drawbacks have motivated research on methods to estimate the AHI from alternative sensor modalities which can be more conveniently acquired, for prolonged periods of time and at home. Photoplethysmography (PPG) is particularly promising in this regard. PPG measures blood volume changes in the microvascular tissue bed and has been extensively used to measure pulse rate and heart rate variability (HRV). In addition, there is a well-described respiratory-induced variation in the PPG signal due to changes in intrathoracic pressure and corresponding changes in cardiac venous return [4], enabling the estimation of respiratory effort [5]. Sudden changes in respiration, e.g. due to deep inspiratory gasps, have also been described, manifesting as abrupt changes in the amplitude of the PPG signal [4].

Most PSG and HSAT setups include PPG in the form of pulse oximetry by transmissive PPG sensors placed on the fingertip to measure peripheral oxygen saturation (SpO2). As such, the suitability of PPG as a single sensor monitoring modality has been the focus of research for a while. Early studies used cardiopulmonary coupling combined with SpO2 [6] and manually engineered features from both signals to detect respiratory events and estimate AHI [7,8]. More recent research has leveraged deep learning, with multitask neural network architectures jointly performing sleep staging and respiratory event detection [9]. However, SpO2 might not always be easy to obtain using simpler, wearable devices. Many devices only contain PPG sensors with single-wavelength LEDs, whereas SpO2 requires multi-wavelength measurements. Moreover, modern reflective PPG sensors that claim to be able to measure SpO2 have only been validated for accuracy in very few studies [10,11], and it remains unclear whether they are sensitive enough to detect brief and mild desaturations, raising questions regarding their clinical applicability in the context of OSA [12].

Given these limitations, we decided to employ reflective wrist-worn single-wavelength PPG without SpO2 as the prime sensor to estimate the AHI. Initial work using manually crafted HRV features extracted from wrist-worn PPG together with accelerometer data, showed promising results in the detection of OSA at different levels of severity [13]. In more recent work, we used neural networks to estimate AHI from cardiorespiratory signals, but still relying on electrocardiography and thoracic RIP belts as input. Together with sleep stages inferred from the same signals, we trained a neural network to detect apnea and hypopnea events. The AHI derived from neural network outputs achieved a high intraclass correlation (ICC) of 0.91 against PSG [14].

In the present study, we developed an algorithm to estimate the AHI based on wrist-worn reflective PPG. We subsequently validated the method against gold-standard PSG on a clinical cohort of patients with multiple sleep disorders, and on an independent, external dataset with participants suspected of OSA.

## 2. Methods

### 2.1 Data

This study employed retrospective data from four distinct datasets. The first two datasets consisted of PSG recordings from the National Sleep Research Resource [15], specifically the Multi-Ethnic Study of Atherosclerosis (MESA) [16] and the Cleveland Family Study (CFS) [17]. These were restricted to adult participants (aged 18 or older) with finger-worn transmissive PPG signals who consented to commercial data use.

The third dataset was a combination of overnight PSGs from the SOMNIA study [18] conducted on unselected adult patients referred for clinical assessment, and the HealthBed study [19], consisting of healthy adult volunteers. Both were recorded at the Sleep Medicine Center Kempenhaeghe and received ethical approval from medical ethical committee at Máxima Medical Center (W17.128, NL63360.015.17). All participants gave informed consent, and the use of these databases for the current study was approved by the medical ethical review board of the Kempenhaeghe hospital (Heeze, the Netherlands). For simplicity, this combined dataset is referred to as “SOMNIA.”

The fourth dataset comprised ambulatory PSGs from the CHARISMA study [20], involving adults suspected of obstructive sleep apnea (OSA) and recorded at the Center for Sleep Medicine Amphia, Breda, The Netherlands. The study was approved by the medical ethical committee of the Máxima Medical Center, Veldhoven, the Netherlands (study number: W20.090) and all participants gave informed consent.

All studies met the ethical principles of the Declaration of Helsinki, the guidelines of Good Clinical Practice and the current legal requirements.

The SOMNIA and CHARISMA datasets included wrist-worn reflective PPG (32 Hz, green LED) and accelerometry (128 Hz) recorded using a watch-like device with a silicon wrist strap.

To ensure consistency across the four datasets, sleep stages and respiratory events were automatically scored using Somnolyzer 4.1 (part of the Sleepware G3 software; Philips RS North America LLC, USA), configured to follow the scoring guidelines from the American Academy of Sleep Medicine (AASM). A validation of the software against manual expert scoring demonstrated agreement non-inferior to human inter-rater reliability [21], and received AASM autoscoring certification [22]. In our prior work using the CFS, MESA, and SOMNIA datasets [14], the Somnolyzer-derived AHI showed excellent agreement with manual scoring (ICC of 0.96), confirming its suitability for this task.

The hypnograms and respiratory events scored by Somnolyzer were used to calculate ground-truth total sleep time (TST_PSG_) and apnea-hypopnea index (AHI_PSG_), and to generate training and validation targets for the model.

### 2.2 PPG-based AHI estimation

Accurate AHI calculation requires estimating the total sleep time and excluding respiratory events detected during wake periods. As detailed in Supplementary Materials, we used two validated sleep staging algorithms to derive a hypnogram and estimate the total sleep time (TST_PPG_). Whenever reflective wrist-PPG and accelerometry were available, we used an algorithm developed for these signals [23]. Otherwise, an algorithm optimized for transmissive finger-PPG [24] was used.

For respiratory event detection, we extended a neural network originally designed for ECG and RIP inputs [14]. We added a PPG encoder to generate a latent representation of the raw PPG data and provided hypnograms (estimated from PPG) as an additional input. The classifier outputs probabilities for four classes at a target sampling rate of 2 Hz: obstructive apnea, central apnea, hypopnea, or normal breathing. The network architecture, comprising 150710 trainable parameters, is illustrated in Figure 1. Contiguous segments where the combined probability for all apneic event types exceeded a threshold determined on the training set, were detected as respiratory events (Figure 2). The AHI_PPG_ was then calculated as the number of detected events during sleep divided by TST_PPG_.

**Figure 1.**
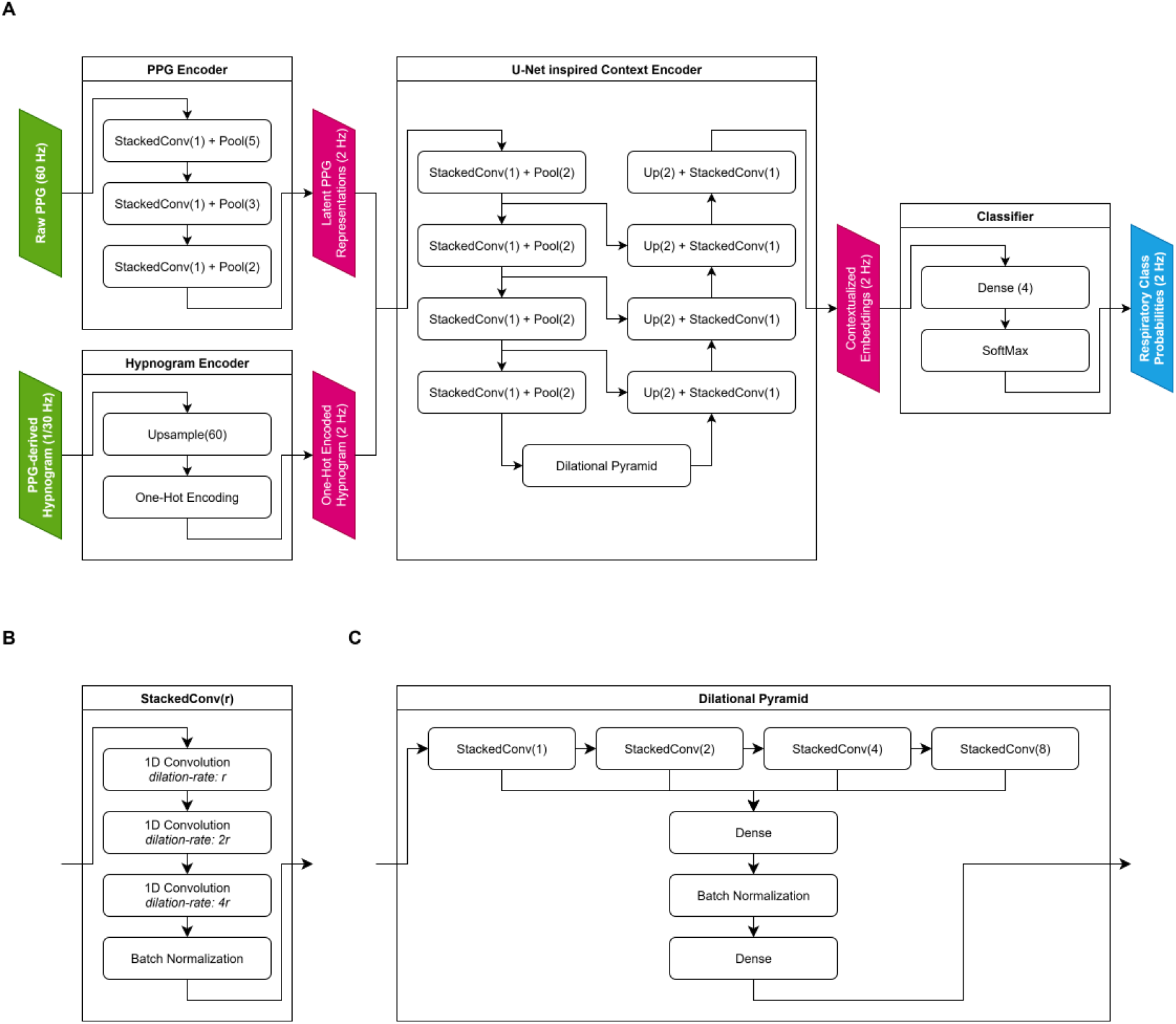
(A) Architecture of the neural network for event detection (B) Architecture of stacked convolution layers used in the model; (C) Architecture of the dilational pyramid used in the context encoder.

**Figure 2.**
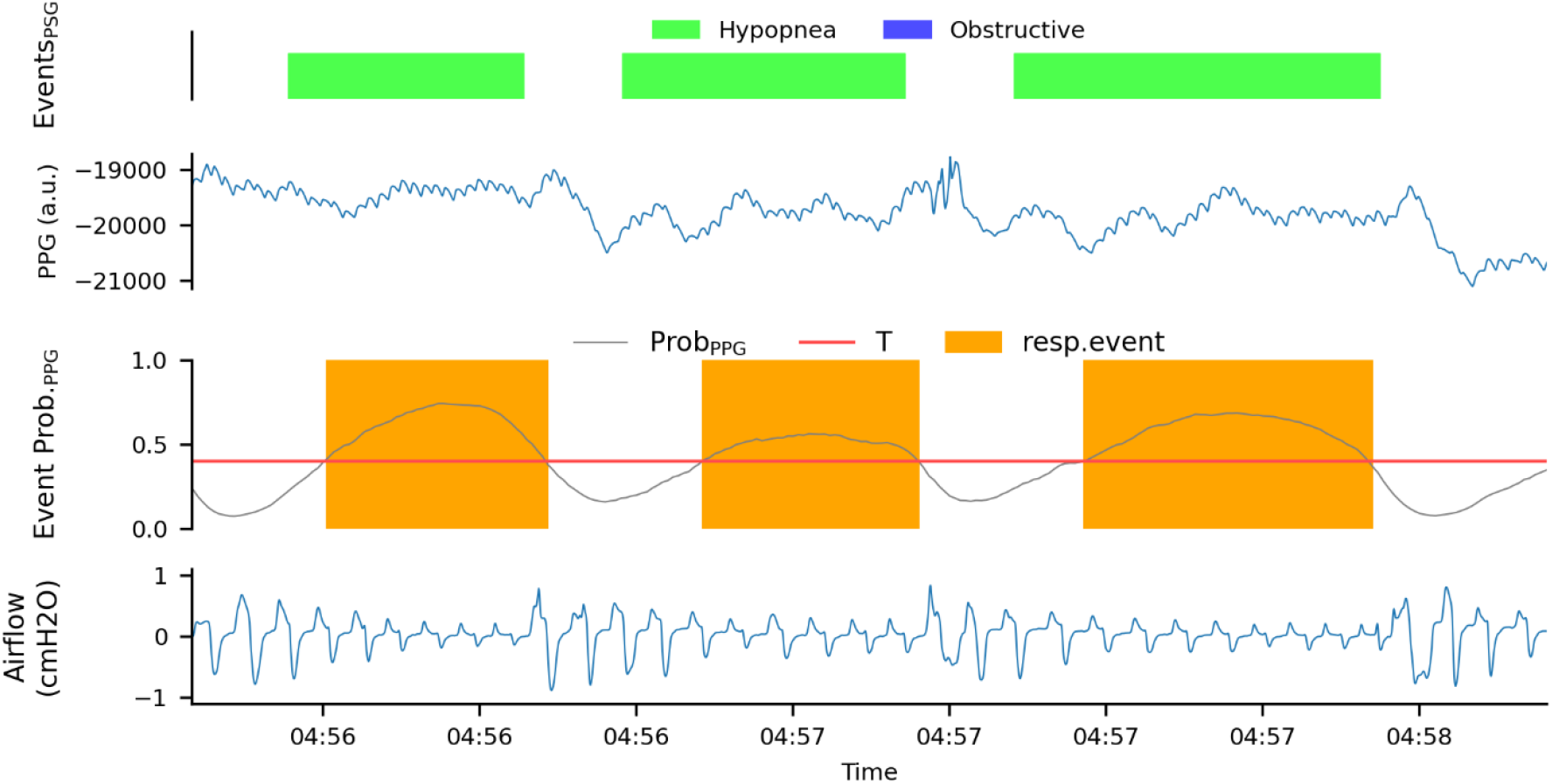
An example period with three hypopneas scored from PSG, and three respiratory events detected from PPG. Large amplitude swings in the PPG follow the termination of each event, coinciding with the hyperpneas evident from the concurrent airflow signal (not used as input to the algorithm) as the events are resolved.

#### 2.2.1 Training

To create the “Development set”, recordings from the MESA, CFS, and SOMNIA databases were grouped into bins based on their AHI_PSG_ values. To preserve the overall AHI distribution across the subsets, participants were randomly sampled at equal proportions into a training set used for model optimization (75%), a validation set used for model selection (4%), and a test set used for internal performance evaluation (21%) (Figure 3). Data splitting was performed at the recording level to ensure that all samples from any given participant were assigned exclusively to one subset (model fitting, model selection, or model testing) and never distributed across multiple subsets. None of the recordings of CHARISMA were used to develop or tune any of the models developed and evaluated in this study. Instead, it was used to perform an external hold-out validation of the final model.

**Figure 3.**
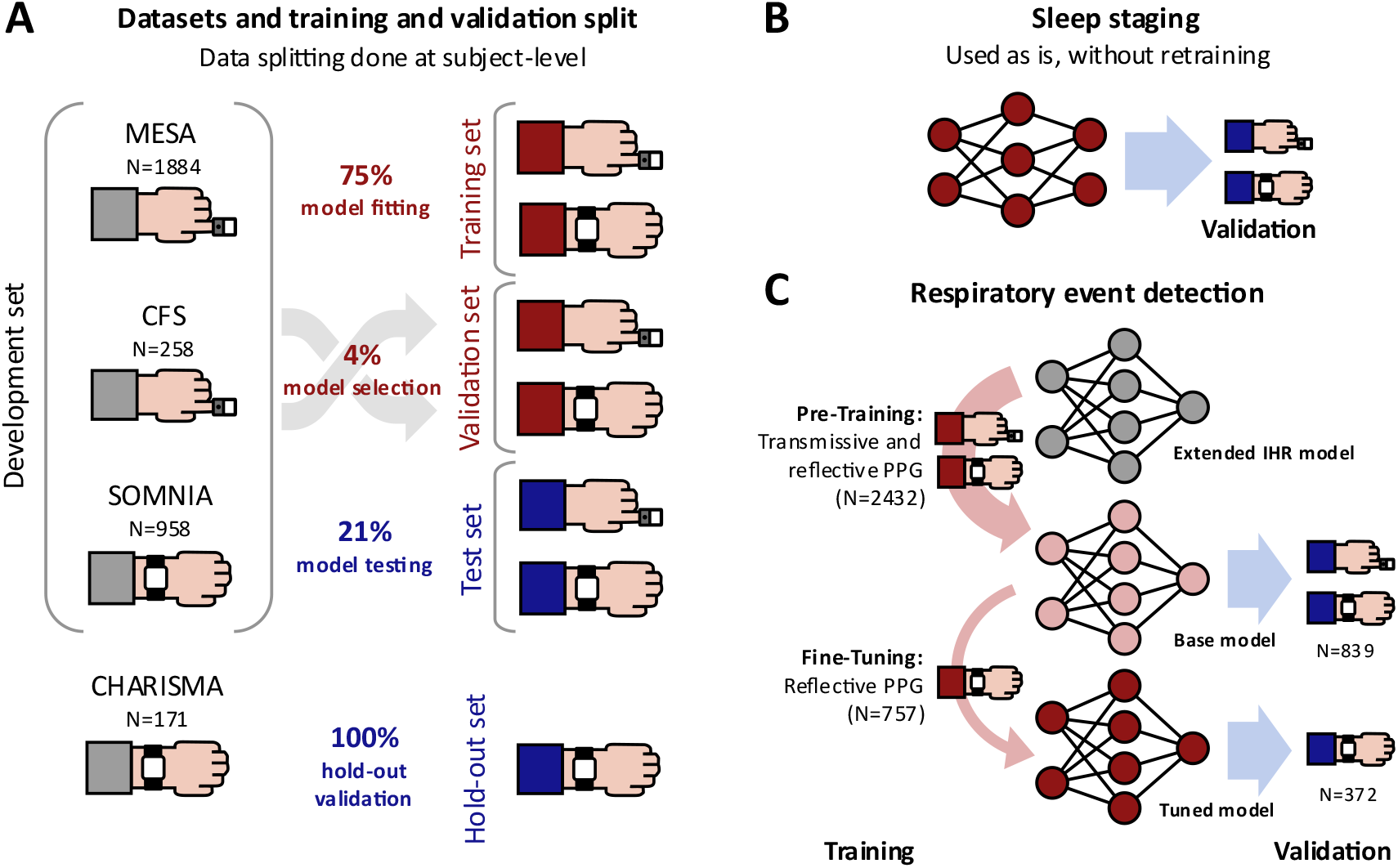
(A) Datasets with concurrent recording of polysomnography and transmissive finger-worn, and reflective wrist-worn photoplethysmography, and training and validation splits; (B) validation of sleep staging neural network; (C) training and validation of sleep respiratory event detection neural network.

We pre-trained a base model able to handle any type of PPG signal as input (transmissive or reflective, finger- or wrist-mounted) using the entire training and validation set. Afterwards, the base model was fine-tuned to the characteristics of reflective wrist-worn PPG signals by restricting the training and validation samples to data from the SOMNIA database. To avoid overfitting on the smaller training set during finetuning, transfer learning [25] was applied to parameters for the PPG encoder and the final stacked convolution, while the weights of the context encoder were frozen, reducing the number of trainable parameters to 18852. An Adam optimizer was used to minimize the cross-entropy classification loss into the four target classes. Training of the base model was stopped after the loss on the validation set did not decrease for 100 consecutive training iterations. Finetuning was performed for 50 training iterations. The thresholds for event detection were identified in a grid-search, maximizing the intraclass correlation coefficient between AHI_PSG_ and the AHI_PPG_ over the combined training set and validation set.

Table 1 indicates the number of recordings of each database and their use in training, tuning and testing the event detection models.

**Table 1.** Number of recordings of each database used for training, validation, testing and hold-out testing of the base and tuned models.

| Dataset | Purpose | Source | PPG Sensor | Model(s) | Recordings |
| --- | --- | --- | --- | --- | --- |
| Training set | Model fitting | CFS | Transmissive | base | 188 |
|  |  | MESA | Transmissive | base | 1402 |
|  |  | SOMNIA | Reflective | base and fine-tuned | 715 |
| Validation set | Model selection<br>(early stopping) | CFS | Transmissive | base | 15 |
|  |  | MESA | Transmissive | base | 70 |
|  |  | SOMNIA | Reflective | base and fine-tuned | 42 |
| Test set | Model testing | CFS | Transmissive | base | 55 |
|  |  | MESA | Transmissive | base | 412 |
|  |  | SOMNIA | Reflective | base and fine-tuned | 201 |
| Hold-out set | Hold-out testing | CHARISMA | Reflective | fine-tuned | 171 |

#### 2.2.2 Validation

Performance was assessed by comparing the AHI_PPG_ and the reference AHI_PSG_ in terms of ICC (two-way random-effects model for absolute agreement) [26]. Performance evaluation was stratified by model (base and fine-tuned), dataset (combined training and validation, test, and hold-out), and input signal type (wrist-worn reflective and transmissive finger PPG). To investigate a possible confounding impact of the performance of PPG-based sleep staging in the estimation of the AHI, we also calculated the ICC between TST_PPG_ and TST_PSG_.

The performance of the fine-tuned model was further evaluated on a combined test set comprising all reflective PPG recordings of the testing set and all recordings of the hold-out set. Besides the ICC with 95% confidence intervals, we also report Spearman’s correlation, root mean square error (RMSE), error estimation bias, standard deviation of the error, 95% limits of agreement (LoA) and a scatter and Bland-Altman analysis of AHI estimation performance.

To assess the added value of using the raw PPG signal instead of PPG-derived instantaneous heart rate (IHR) as input, we repeated the main experiment replacing the latent representations of the raw PPG signal with PPG-derived IHR. We further evaluated the added value of the two-step training procedure by omitting the pre-training step and instead training the entire model purely on reflective wrist-PPG data used for fine-tuning in the original experiment. To ensure comparable results, we used the same data splits in all experiments.

#### 2.2.3 OSA severity classification and diagnostic performance

To assess OSA severity classification performance, we calculated a confusion matrix using the canonical thresholds of 5, 15, and 30 to separate four severity classes: no (OSA), mild, moderate, and severe OSA. To account for uncertainty around the boundaries and to avoid an overly pessimistic view of performance due to small errors that might not be clinically relevant, we performed the same severity classification evaluations using a near-boundary double-labelling (NBDL) procedure [27] defining near-boundary zones for the reference AHI_PSG_ of 2.4 ≤ AHI < 7.0 for ‘no OR mild OSA’, 12.4 ≤ AHI < 17.4 for ‘mild OR moderate OSA’, and 26.6 ≤ AHI < 35.2 for ‘moderate OR severe OSA’.

We reported the accuracy, sensitivity, specificity, positive predictive value (PPV), negative predictive value (NPV), positive and negative likelihood ratios (LR+, LR-) for three binary tasks, classifying participants with at least mild (versus no OSA), moderate (versus no OSA or mild OSA), or severe OSA (versus no OSA, or lower severities) based on canonical thresholds as well as NBDL.

Throughout the study we used a Shapiro-Wilk normality test to determine whether the sample data was normally distributed (at a p-value of 0.05). Since nearly none of the data analyzed was normally distributed, all reported statistics indicate the median, and between curly brackets, the 25th and 75th percentiles: “median {Q1, Q3}”.

## 3. Results

### 3.1 Demographics

Table 2 shows the demographic characteristics of subjects in each dataset, including the type of PPG signal available.

**Table 2.**
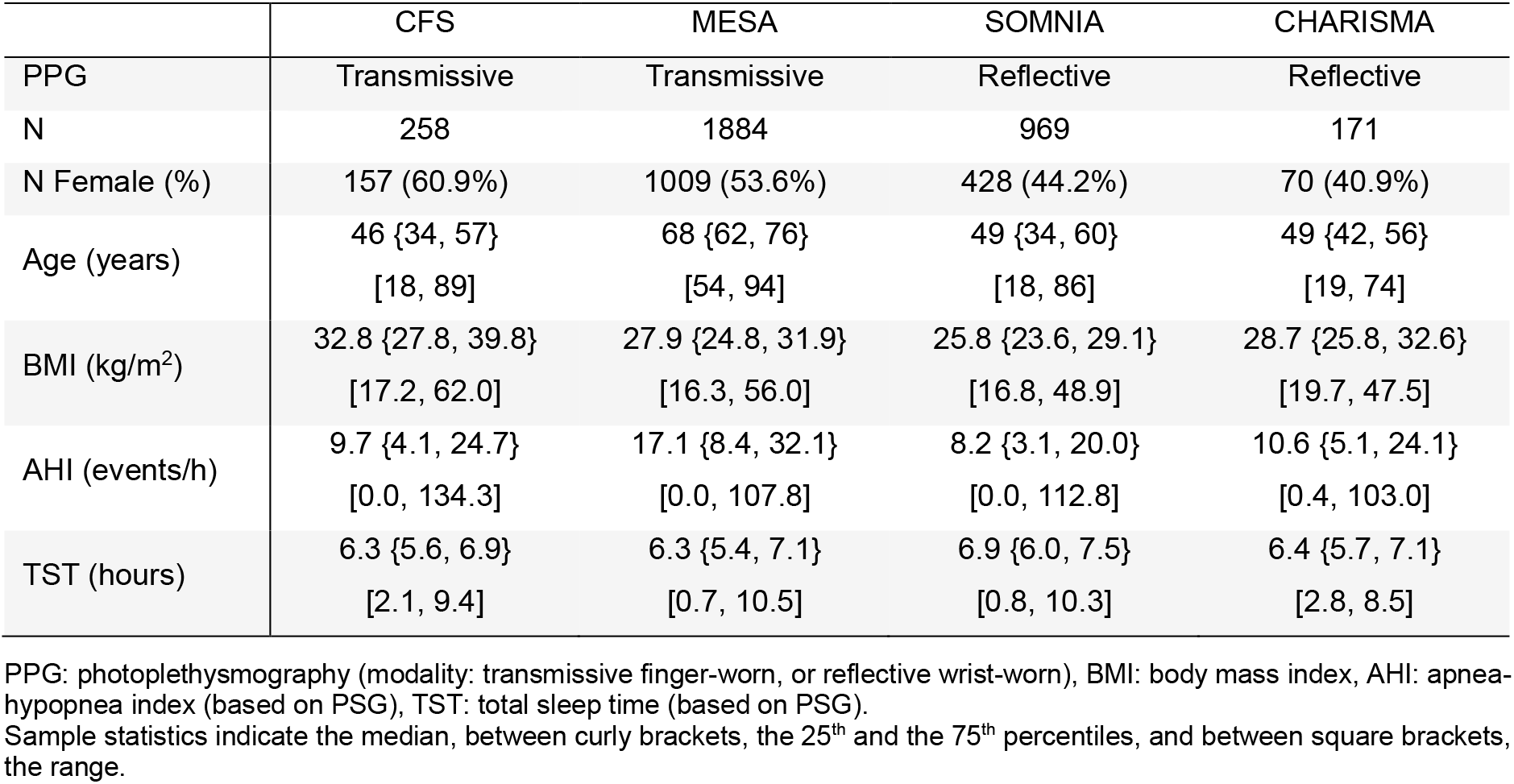
Participant demographics of each dataset.

### 3.2 AHI estimation performance

The base model was trained for a total of 197 iterations and after optimization on the training set, we obtained a threshold for the respiratory event probabilities of 0.45. The fine-tuned model was trained for a fixed total of 50 iterations, and we obtained a probability threshold of 0.40.

Table 3 shows the agreement for the estimated AHI and TST compared to the corresponding ground-truth obtained from PSG. The performance of AHI estimation with the fine-tuned model consistently surpassed that of the base model, both in the reflective PPG recordings of the testing split of the development set (SOMNIA) and in the hold-out set (CHARISMA). Additionally, there was a notable difference in TST estimation performance between the transmissive and reflective recordings of the development set, with ICCs of 0.78 and 0.80 in the transmissive recordings, and 0.87 and 0.88 in the reflective recordings of the training and testing splits, respectively.

**Table 3.** Intraclass correlation coefficient for AHI and TST estimation based on PPG in comparison with the reference ground-truth obtained from PSG for the base and fine-tuned models, for the training, testing and hold-out validation sets.

| Dataset | Signal | Sources | N | TST | AHI |  |
| --- | --- | --- | --- | --- | --- | --- |
|  |  |  |  |  | Base | Fine-tuned |
| Training | Combined | CFS, MESA, SOMNIA | 2432 | 0.81<br>[0.79, 0.83]* | 0.94<br>[0.93, 0.94]* | - |
|  | Transmissive only | CFS, MESA | 1675 | 0.78<br>[0.76, 0.80]* | 0.94<br>[0.93, 0.95]* | - |
|  | Reflective only | SOMNIA | 757 | 0.87<br>[0.76, 0.92]* | 0.91<br>[0.86, 0.94]* | 0.95<br>[0.94, 0.96]* |
| Testing | Combined | CFS, MESA, SOMNIA | 668 | 0.83<br>[0.80, 0.85]* | 0.91<br>[0.90, 0.93]* | - |
|  | Transmissive only | CFS, MESA | 467 | 0.80<br>[0.77, 0.83]* | 0.93<br>[0.91, 0.94]* | - |
|  | Reflective only | SOMNIA | 201 | 0.88<br>[0.79, 0.93]* | 0.84<br>[0.76, 0.88]* | 0.87<br>[0.83, 0.90]* |
| Hold-Out | Reflective only | CHARISMA | 171 | 0.88<br>[0.68, 0.94]* | 0.90<br>[0.86, 0.93]* | 0.91<br>[0.88, 0.93]* |

ICC: intraclass correlation (two-way random-effects model for absolute agreement); AHI: apnea-hypopnea index; TST: total sleep time
Both the AHI and the TST rows indicate the intra-class correlation coefficient (two-way random-effects model for absolute agreement) and 95% confidence intervals (\* $p < 0.001$ ).
The overall performance of the 'base model' on the left-most column corresponds to the aggregation of the performance of that model for the recordings with transmissive and with reflective PPG in the training and in the testing sets. Also, note that the performance for TST is the same with the base since they are based on the same sleep stage classification algorithm.
Note that there is no difference in performance for TST with the base model (reflective) and the tune model, since the PPG- (and accelerometer) based sleep staging algorithm was used as is, without retraining or tuning.

Focusing only on the fine-tuned model, Table 4 indicates the combined performance on the test set recordings of SOMNIA and the recordings of the hold-out set CHARISMA. Overall, the algorithm achieved a Spearman’s correlation between AHI_PPG_ and AHI_PSG_ of 0.83, and an overall ICC of 0.89. The RMSE, bias, and 95% limits of agreement (LoA) were almost the same for the hold-out set (CHARISMA) as for the testing split of the development set (SOMNIA). Figure 4 illustrates the scatter and Bland-Altman plots for AHI estimation. Bias and 95% LoA were nearly the same for both test datasets. There was no evidence of proportional bias, but we noted some heteroscedasticity for values up to 15, where the variance in the estimation error increased proportionally with the AHI.

**Table 4.**
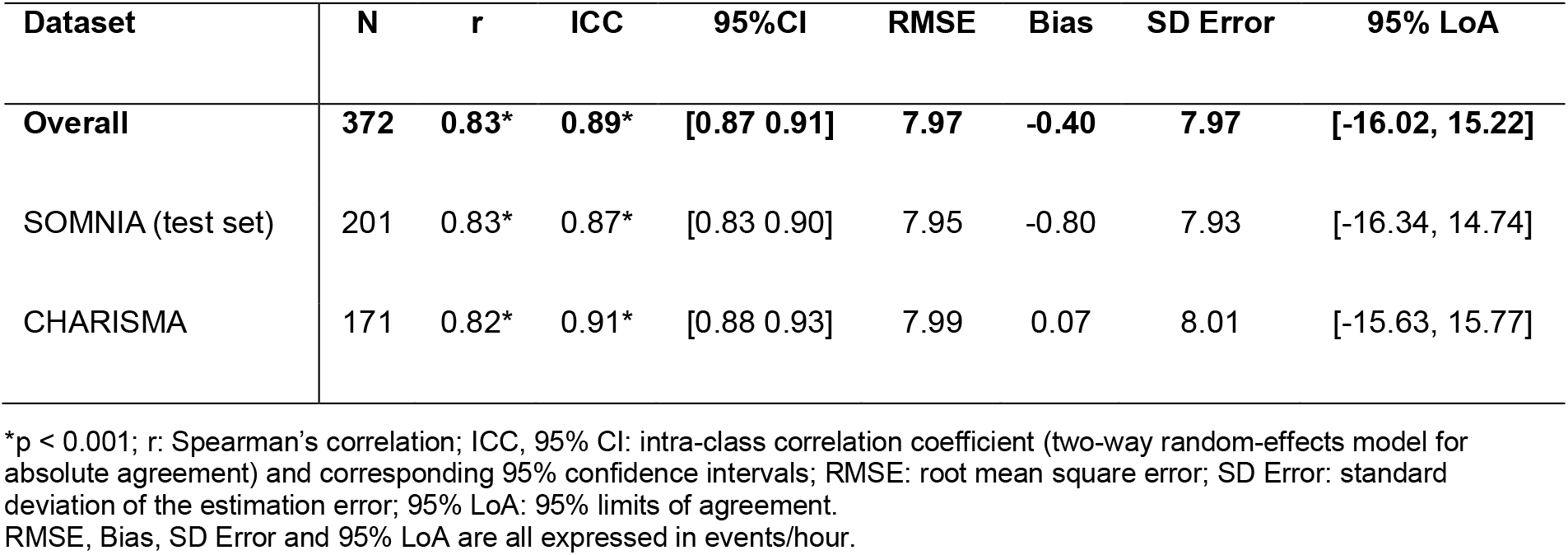
Combined AHI estimation performance with the fine-tuned model on the test set (reflective wrist-PPG recordings only) and hold-out set.

| Dataset | N | r | ICC | 95%CI | RMSE | Bias | SD Error | 95% LoA |
| --- | --- | --- | --- | --- | --- | --- | --- | --- |
| Overall | 372 | 0.83* | 0.89* | [0.87 0.91] | 7.97 | -0.40 | 7.97 | [-16.02, 15.22] |
| SOMNIA (test set) | 201 | 0.83* | 0.87* | [0.83 0.90] | 7.95 | -0.80 | 7.93 | [-16.34, 14.74] |
| CHARISMA | 171 | 0.82* | 0.91* | [0.88 0.93] | 7.99 | 0.07 | 8.01 | [-15.63, 15.77] |

\* $p < 0.001$ ; r: Spearman's correlation; ICC, 95% CI: intra-class correlation coefficient (two-way random-effects model for absolute agreement) and corresponding 95% confidence intervals; RMSE: root mean square error; SD Error: standard deviation of the estimation error; 95% LoA: 95% limits of agreement. RMSE, Bias, SD Error and 95% LoA are all expressed in events/hour.

**Figure 4.**
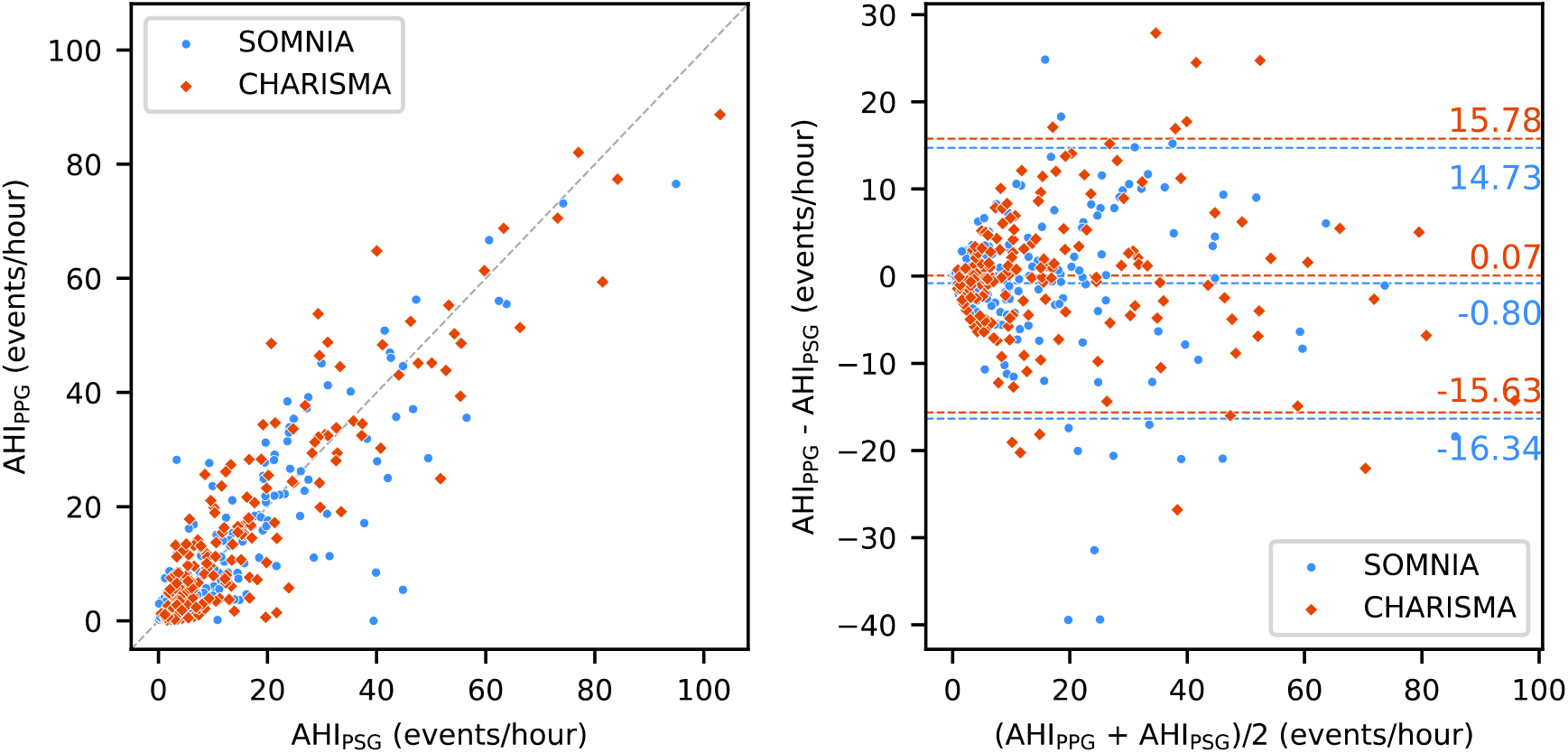
(Left) scatter and (right) Bland-Altman plots for AHI estimated from reflective wrist-PPG against reference ground-truth AHI obtained from PSG for all test set recordings of SOMNIA, and all recordings of CHARISMA. The horizontal dashed lines in the Bland-Altman plot indicate the bias and 95% limits of agreement for each separate dataset.

In the second experiment, we used PPG-derived IHR (together with sleep stages) as input to the respiratory event detection algorithm, and obtained, on the same 668 recordings of the test portion of the development set, an ICC of 0.80 (95% CI [0.75, 0.84]). Restricting the evaluation to the 201 recordings with reflective wrist-worn PPG of the test portion of the development set, we obtained an ICC of 0.77 (95% CI [0.71, 0.82]). This performance was thus substantially lower than the ICC of 0.91 and 0.84 obtained on the same recordings with the base model in the original experiment using the raw PPG signal as input.

Finally, in the third experiment we used only reflective-PPG recordings to train the neural network and obtained an ICC of 0.76 (95% CI [0.69 0.81]) on the same 201 SOMNIA recordings of the testing split of the development set. This performance was substantially lower than the ICC of 0.84 and 0.87 from the original experiment with the base model and the fine-tuned model, respectively.

### 3.3 OSA severity classification performance

Table 5 (left) indicates the confusion matrix for OSA severity classification on the combined test and hold-out sets with four classes using the canonical OSA severity thresholds, achieving a classification accuracy of 65.6% and a Cohen’s kappa coefficient of agreement of 0.53. Using NBDL (Table 5, right) the classification accuracy increased to 84.1% and Cohen’s kappa to 0.78. Notably, only nine participants were erroneously classified with more than one severity class away from the ground-truth using either evaluation method.

**Table 5.**
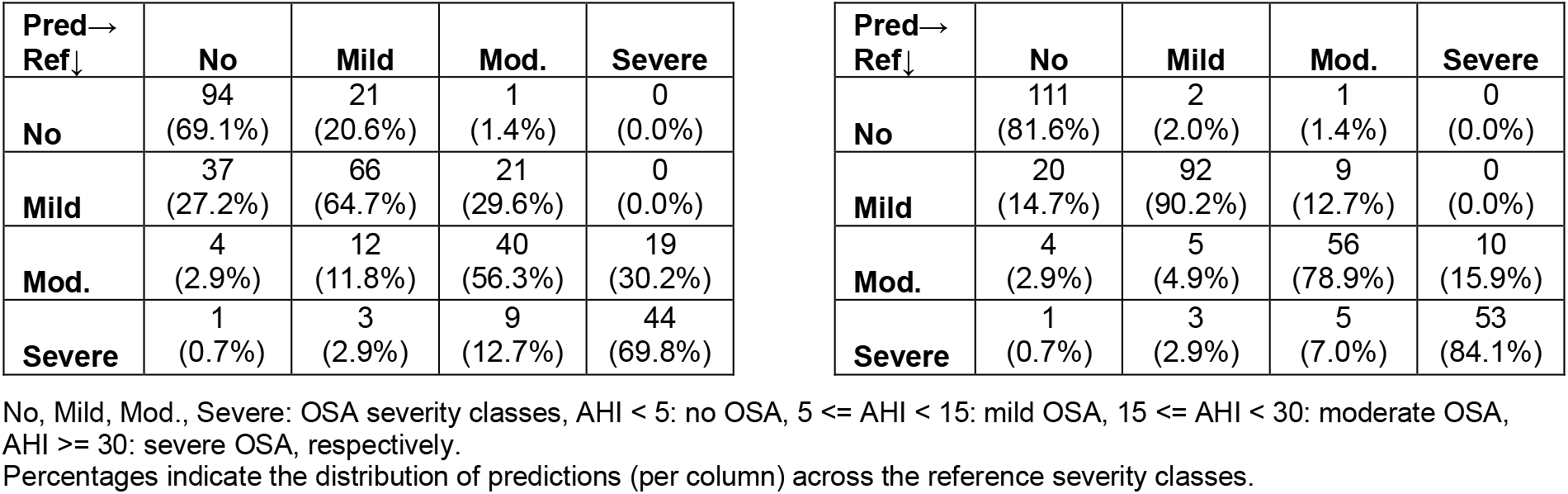
Confusion matrix for OSA severity classification with (left) canonical thresholds and (right) with near-boundary double labelling.

Table 6 indicates the diagnostic performance for OSA severity estimation with different thresholds, using canonical thresholds and NBDL. Performance across all metrics is always higher than 80%, except for PPV for severe OSA detection, with 69.8%, and NPV for detection of OSA (of any severity), at 69.1%.

**Table 6.** Diagnostic performance for canonical and near-boundary double labelling.

| Min. Severity | Prevalence (%) | Accuracy | Sensitivity | Specificity | PPV | NPV | LR+ | LR- |
| --- | --- | --- | --- | --- | --- | --- | --- | --- |
| **Canonical thresholds** |  |  |  |  |  |  |  |  |
| Mild (AHI ≥ 5) | 68.8 | 0.828 | 0.836 | 0.810 | 0.907 | 0.691 | 4.41 | 0.202 |
| Moderate (AHI ≥ 15) | 35.5 | 0.887 | 0.848 | 0.908 | 0.836 | 0.916 | 9.26 | 0.167 |
| Severe (AHI ≥ 30) | 15.3 | 0.914 | 0.772 | 0.940 | 0.698 | 0.958 | 12.8 | 0.243 |
| **Near-boundary double labelling** |  |  |  |  |  |  |  |  |
| Mild (AHI ≥ 5) | 69.4 | 0.925 | 0.903 | 0.974 | 0.816 | 0.987 | 34.3 | 0.100 |
| Moderate (AHI ≥ 15) | 36.8 | 0.938 | 0.905 | 0.957 | 0.945 | 0.925 | 21.3 | 0.099 |
| Severe (AHI ≥ 30) | 16.7 | 0.949 | 0.855 | 0.968 | 0.971 | 0.841 | 26.5 | 0.150 |

Min. Severity: minimum severity threshold defined for positive class (i.e., positive class includes this severity and higher) for each binary classification task using canonical thresholds and using near-boundary double-labelling.
Prevalence: percentage of participants with severity (based on AHI from PSG) equal or greater than each severity threshold; PPV: positive predictive value, NPV: negative predictive value, LR+, LR-: positive and negative likelihood ratios.

## 4. Discussion

We demonstrated the feasibility of estimating the AHI using only single-wavelength reflective wrist-worn PPG. In comparison with ground-truth AHI derived from PSG, the method achieved performance comparable to state-of-the-art algorithms relying on more direct assessment of cardiac and respiratory signals. The testing split of the development dataset included both healthy participants and individuals with a range of sleep disorders including, but not limited to, OSA. In this dataset, the model achieved an ICC of 0.87. External validation on an independent cohort referred for PSG due to suspected OSA yielded an ICC of 0.91. Across both datasets, this corresponded to a combined ICC of 0.89, with a negligible mean bias of –0.40 events/hour and an RMSE of 7.97 events/hour.

Using only a single-sensor input for respiratory event detection, the algorithm matched the performance demonstrated in previous approaches that relied on both ECG and respiratory belt inputs. This adaptation highlights two key insights. First, the addition of dilated convolutions enabled the network to replace the two original inputs (ECG-derived heart rate and respiratory effort from RIP) with a single PPG signal, eliminating the need for beat detection. Second, the modified network effectively leverages respiratory information directly from raw PPG data. As shown in Figure 2, PPG amplitude swings frequently accompany respiratory events, reflecting intrathoracic pressure changes during the events and as they resolve. These swings have been used to estimate respiratory effort from PPG [5], but also described as artifacts from abrupt respiratory changes, such as deep gasps [4]. This likely explains the algorithm’s sustained high performance, even without a dedicated respiratory effort input, and the superior results compared to using only PPG-derived instantaneous heart rate.

Several studies have used finger PPG to estimate AHI. Using PPG and SpO_2_, Hilmisson et al. reported a Pearson correlation of 0.95 with the PSG-derived AHI in a pediatric population of 805 children [6]. Lazazzera et al. reported a correlation of 0.88 with PSG in a cohort of 96 adults suspected of obstructive sleep apnea (OSA) [7]. Using a neural network model using PPG and SpO2, Huttunen et al. reported an intraclass correlation coefficient (ICC) of 0.946 in a test set of 88 participants with and without OSA [9]. Finally, Cajal et al. used PPG-derived pulse rate variability and SpO2 features achieving a Pearson’s correlation of 0.89 with PSG-derived AHI in 96 subjects suspected of OSA [8]. Although direct comparisons are limited due to the nature of the different datasets, our approach delivers performance comparable to these other studies but using only single-wavelength reflective PPG.

An essential component in the presented approach is the use of sleep stages, used not only as input to respiratory event detection but also to calculate total sleep time (TST), required to determine the AHI. Although TST estimation was slightly less accurate with transmissive PPG (ICC of 0.80) compared to reflective PPG plus accelerometry (0.88), this did not impact AHI estimation accuracy. In fact, the base model achieved a higher ICC in the transmissive-PPG subset (0.93) than in the reflective-PPG subset (0.84), possibly reflecting the different characteristics of both the sensor and the cohorts.

Our classifier showed consistent performance across datasets. The ICC was slightly higher, and RMSE, bias, and limits of agreement were nearly identical between the hold-out set and the development test split. This suggests strong generalization to new (external) data, including recordings in different environments (ambulatory versus in-lab) and different PSG setups. The slightly lower ICC in the SOMNIA cohort may be due to its clinical diversity, with many participants having comorbid sleep disorders alongside OSA.

Training with reflective wrist-worn PPG alone led to a lower performance than training with both reflective/wrist- and transmissive/finger-PPG, even without transfer learning. This suggests that the additional recordings with transmissive PPG greatly enhanced the overall training process. The increased robustness of the classifier could be caused by the larger number of subjects in the training data, and by different characteristics of the PPG sensors used.

A limitation of the approach remains the lack of SpO2 data, which is not available (or sufficiently accurate) in many wearable PPG devices. This prevents the characterization of desaturation severity and frequency, markers that may be better linked than AHI to some clinical outcomes including daytime symptoms [28] and cardiovascular risk [29]. It may also partly hinder detection of hypopneas. Hypopneas with arousals may be detectable due to the autonomic (arousal) responses visible in PPG [3], but hypopneas only characterized by desaturations are more likely to be missed. The availability of an accurate SpO2 sensor could address this in future work. The validity of the method in the presence of severe cardiac arrhythmia and under medication altering autonomic nervous system activity remains to be investigated.

In conclusion, this study demonstrates the use of wrist-worn reflective PPG to obtain the AHI as an indicator of OSA severity, achieving a high estimation performance in comparison with gold standard PSG. This approach enables the use of relatively comfortable sensors integrated into convenient wearables to enable long-term monitoring of sleep-disordered breathing, both during diagnostic assessment and therapy follow-up.

## Data Availability

The CFS and MESA data used in the study are publicly available via the National Sleep Research Resource. The SOMNIA and CHARISMA datasets are not publicly available due to privacy and legal restrictions. Any inquiries should be directed to the corresponding author.

https://sleepdata.org/datasets/mesa

https://sleepdata.org/datasets/cfs

## Ethics approval

The SOMNIA and Healthbed studies received ethical approval from the Máxima Medical Center (W17.128, NL63360.015.17). All participants gave informed consent, and the use of these databases in the current study was approved by the Medical Ethical Committee of the Kempenhaeghe hospital (Heeze, the Netherlands).

The CHARISMA study was approved by the medical ethical committee of the Máxima Medical Center, Veldhoven, the Netherlands (study number: W20.090) and all participants gave informed consent.

## Acknowledgments

The authors would like to thank all somnologists, sleep technicians, and nurses at the Center for Sleep Medicine Kempenhaeghe and at the Amphia Hospital for their vital role in enabling this study.

## Author contributions

PF: Conceptualization, methodology, formal analysis, software, writing – original draft MR: software, writing – original draft, F.v.M.: data curation, writing – reviewing and editing, J.A.: investigation, writing – reviewing and editing, M.v.G.: writing – reviewing and editing, S.O.: investigation, methodology, supervision, writing – reviewing and editing.

