## Supplementary Materials for "Apnea-hypopnea index estimation with wrist-worn photoplethysmography"

### PPG-based sleep staging

In this study, we used two previously developed cardiac-based sleep stage classification algorithms, both of which have been validated on a large group of patients with and without sleep disorders.

The first algorithm is a neural network comprising convolutional and recurrent layers, originally designed to utilize any combination of the following inputs: instantaneous heart rate (IHR) measured with any cardiac sensor, including PPG; respiratory airflow; and respiratory effort measured with thoracic or abdominal belts [1]. The algorithm outputs a classification for each 30-second epoch into one of four classes: Wake, combined N1-N2, N3, and REM. We used this same algorithm in our previous work on AHI estimation with ECG and respiratory effort [2], with the key differences that in the present study we used IHR derived from PPG (instead of ECG) and did not use any of the respiratory inputs (whereas in the previous study we used respiratory effort). The algorithm was obtained and used as is, without retraining or tuning to the characteristics of the current study population.

The second algorithm was originally developed to perform automatic sleep staging based on sensor signals obtained from wrist-mounted devices. Namely, it was trained to use IHR, obtained from reflective PPG, and activity counts, obtained from an accelerometer [3]. This algorithm also outputs four classes, and was also used as is, without retraining to the current study population.

In all recordings, the PPG signal (transmissive and reflective) was first pre-processed to obtain a time series of IHR, required by both sleep staging algorithms. This was achieved by first detecting the location of individual beats (pulses) on the PPG signal using a previously described algorithm [4]. In short, pulses were detected as the points where the first derivative of the signal crosses zero towards a positive value, corresponding to the throughs in the waveform. The interval between consecutive fiducial points was used to represent a time series with inter beat intervals (IBI) which was finally sampled at 10 Hz, inverted and multiplied by 60 to obtain a representation of IHR in beats per minute (bpm).

Since an accelerometer input was not available in the datasets with transmissive PPG (MESA and CFS), sleep staging was performed for the recordings in both datasets using the first algorithm.

In recordings where an accelerometer was available (SOMNIA, CHARISMA), the second algorithm was used, which required a second input to the sleep staging algorithm, namely a gross measure of body movements, or activity counts. This was computed as described in earlier work [5] and summarized here. Activity counts were calculated for each 30 second epoch using the three-axis accelerometer signal from the wrist-worn device available in these datasets. A low-pass filter with a third-order Butterworth filter and a 1 Hz cutoff frequency was first applied on each accelerometer axis. For each non-overlapping 1-second window, a simple gravity estimate was determined by averaging the amplitude of each axis. The activity count for each 1-second interval was then computed by summing the absolute values of each axis and subtracting the gravity estimate for that window. To represent activity counts for each 30-second epoch, the 1-second values within each 30-second window were summed.

In recordings including PPG from a wrist-worn sensor (SOMNIA, CHARISMA) and before performing sleep staging, we synchronized the clock of the PPG and accelerometer with the PSG clock of each recording using the ECG signal recorded during the PSG acquisition. First, we calculated IBI time series from the ECG signal in the PSG recording using a QRS detector, followed by a post-processing localization algorithm [6]. The clock offset and drift of each recording were then determined by maximizing the correlation between the ECG-derived IBI series and the previously described PPG-derived IBI series.

After synchronizing the clocks, and to allow a direct comparison with the PSG scorings of sleep and respiratory events, we limited the PPG and accelerometer (where available) signals to the period coinciding with the “lights off” interval, as annotated in the PSG recordings.

Finally, after obtaining the sleep stages for the lights off period of each recording, we calculated the total sleep time, $TST_{PPG}$, by summing the time on epochs of any sleep stage (i.e., except Wake) as scored by either of the two algorithms.
